# The Project for Objective Measures Using Computational Psychiatry Technology (PROMPT): Rationale, Design, and Methodology

**DOI:** 10.1101/19013011

**Authors:** Taishiro Kishimoto, Akihiro Takamiya, Kuo-ching Liang, Kei Funaki, Takanori Fujita, Momoko Kitazawa, Michitaka Yoshimura, Yuki Tazawa, Toshiro Horigome, Yoko Eguchi, Toshiaki Kikuchi, Masayuki Tomita, Shogyoku Bun, Junichi Murakami, Brian Sumali, Tifani Warnita, Aiko Kishi, Mizuki Yotsui, Hiroyoshi Toyoshiba, Yasue Mitsukura, Koichi Shinoda, Yasubumi Sakakibara, Masaru Mimura, on behalf of the PROMPT collaborators

**Author notes:** **Corresponding Author** Taishiro Kishimoto, MD, PhD, Associate Professor of Psychiatry, Keio University School of Medicine, Tokyo, Japan.

## Abstract

**Background:** Depressive and neurocognitive disorders are debilitating conditions that account for the leading causes of years lived with disability worldwide. Overcoming these disorders is an extremely important public health problem today. However, there are no biomarkers that are objective or easy-to-obtain in daily clinical practice, which leads to difficulties in assessing treatment response and developing new drugs. Due to advances in technology, it has become possible to quantify important features that clinicians perceive as reflective of disorder severity. Such features include facial expressions, phonic/speech information, body motion, daily activity, and sleep. The overall goal of this proposed study, the Project for Objective Measures Using Computational Psychiatry Technology (PROMPT), is to develop objective, noninvasive, and easy-to-use biomarkers for assessing the severity of depressive and neurocognitive disorders.

**Methods:** This is a multi-center prospective study. DSM-5 criteria for major depressive disorder, bipolar disorder, and major and minor neurocognitive disorders are inclusion criteria for the depressive and neurocognitive disorder samples. Healthy samples are confirmed to have no history of psychiatric disorders by Mini-International Neuropsychiatric Interview, and have no current cognitive decline based on the Mini Mental State Examination. Participants go through approximately 10-minute interviews with a psychiatrist/psychologist, where participants talk about non-specific topics such as everyday living, symptoms of disease, hobbies, etc. Interviews are recorded using RGB and infrared cameras, and an array microphone. As an option, participants are asked to wear wrist-band type devices during the observational period. The interviews take place ≤10 times within up to five years of follow-up. Various software is used to process the raw video, voice, infrared, and wearable device data. A machine learning approach is used to predict the presence of symptoms, severity, and the improvement/deterioration of symptoms.

**Discussion:** The PROMPT goal is to develop objective digital biomarkers for assessing the severity of depressive and neurocognitive disorders in the hopes of guiding decision-making in clinical settings as well as reducing the risk of clinical trial failure. Challenges may include the large variability of samples, which makes it difficult to extract the features that commonly reflect disorder severity.

**Trial Registration:** UMIN000021396, University Hospital Medical Information Network (UMIN)

## Background

Depressive disorders and neurocognitive disorders are common, disabling, and debilitating psychiatric conditions. Major depressive disorder (MDD) affects approximately 6% of the adult population worldwide each year [1], and the prevalence in 2017 is estimated to have been 264.5 million people [95% uncertainty interval (UI) 246.3 to 286.3]. Moreover, depressive disorder is the third leading cause of years lived with disability (YLDs) that contributes to 43.1 million YLDs (95%UI 30.5 to 58.9) [2]. Pharmacotherapy is one of the mainstays of depression treatment, and many efforts to develop new antidepressant treatments have been made. However, clinical trials for antidepressant medications face tremendous difficulties. Failures in such clinical trials have even led to the unfortunate consequence of several pharmacological companies moving out of the psychiatric field [3, 4]. The reasons for these clinical trial failures may include multiple factors, such as: 1) the mechanisms of an illness are not fully understood; 2) the heterogeneity of the targeted population; 3) difficulty in recruiting patients with severe symptoms; 4) too many placebo responders; and so on. Poor reliability of measurement, poor interview quality, and rater bias are also important factors that contribute to a number of these reasons for trial failure [5, 6]. The most popular severity measurement tools for depression include the Hamilton Depression Rating Scale (HAM-D) [7] and Montgomery-Asberg Depression Rating Scale (MADRS) [8]. Although HAM-D and MADRS are clinician-rated assessment tools, these measures mainly depend on subjective reports by the patients. Such rating scales that rely on patients’ subjective feelings can be influenced by the patient’s personality and/or the interviewer’s ability/skill. It is also common for the anchor point to be ambiguous, among other issues. On the other hand, there are evaluation items in these rating scales that do not rely on the patient’s subjective opinion, such as psychomotor disturbances (i.e., retardation and agitation) in HAM-D, and apparent sadness in MADRS. But even these items still depend on subjective assessments by the clinicians, and are therefore not truly objective. Several other biological, objective methods have been investigated with the aim of ensuring a more objective measurement of depression severity, such as monoamine levels in cerebrospinal fluids [9], cytokines [10], positron emission tomography (PET) [11], neuroendocrine tests [12], and magnetic resonance imaging (MRI) [13]. However, no objective biomarkers that are reliable and easy-to-use in clinical settings have been discovered.

Dementia, another disease targeted by this research, is increasingly affecting people as the global population ages. The number of individuals who live with neurocognitive disorders world-wide is estimated to be 45 million (95%UI 39.7 to 50.4) [2], and these disorders contribute to 6.5 million YLDs (95%UI 4.7 to 8.6). Furthermore, neurocognitive disorders are the fifth leading cause of death globally, accounting for 2.4 million (95% UI 2.1 to 2.8) deaths [14]. It is believed that in the future, this number may increase to up to 82 million by 2030, and 152 million by 2050 [15]. Additionally, mild cognitive impairment (MCI), which is an intermediate stage between the expected cognitive decline of normal aging and the decline caused by a neurocognitive disorder, has an estimated prevalence of 10%-20% in individuals aged ≥65 years [16]. The biological mechanisms of dementia may be better understood than those of depression. Based on the current understanding of those mechanisms, several early diagnosis methods have already been introduced in clinical settings, or have become possible at the research stage; for example, PET imaging, cerebrospinal fluid, and plasma for amyloid-β and/or tau protein detection [17–20]. However, unfortunately, the possibility of using these methods in screenings or illness evaluations is still far off, as these biomarkers do not necessarily reflect real-time cognitive decline, and the examinations required are costly or invasive such that they cannot be repeated well. Moreover, a biomarker may no longer play its role as a clinical marker that reflects symptom severity when the marker itself is the target of the medication, such as amyloid-β or tau protein. Over 100 clinical drug trials targeting dementia have ended in failure, and currently, as treatment practices shift focus to target the very early stage of the illness [21], it is beneficial to develop repeatable tests for discovering preclinical conditions or MCI. There are several rating scales being used in clinical settings to test cognitive function; for example, Mini-Mental State Examination (MMSE) [22] and Montreal Cognitive Assessment (MoCA) [23] are ones widely used around the world. However, these evaluations require testing the subject’s calculation and memorization abilities, which may place an extra mental burden on the subject. Additionally, the subject’s education history can greatly influence the results of these rating scales, and scores can be affected by the tester’s ability/skill. The ceiling effect and floor effect are also issues, and the learning effect is most likely a large problem as well. This is because, as previously stated, as treatment practices shift focus to early detection, patients with slight cognitive impairment may end up memorizing the testing procedures, which would defeat the purpose of the exams.

So far, it has been explained that there are limits to the “gold standard” rating scales used in clinical settings and trials, and that there are no ideal biomarkers. But at the same time, psychiatrists are able to infer a certain amount about a patient’s severity by the way they act in clinical settings; for example, how the patient enters the room, sits in a chair, or speaks to the interviewer. In this way, psychiatrists can observe the patient’s condition and determine if their treatment is effective. In terms of depression, the various domains of human expression, such as facial movements, speech, and motor movements, have been identified as observable features in depressed patients since Hippocrates’s era [24]. Several studies linked depression with less eye contact, overall sluggishness, slumping back posture, etc. [25–28]. These observable psychomotor abnormalities continue to be regarded by experts as essential and critical features of depression, especially melancholic depression or melancholia [29–31]. In regard to neurocognitive disorders, clinicians can gain information from instances when patients hesitate in their speech trying to recall a word, or when they try to gloss over the fact they cannot remember something. As dementia symptoms progress, patients lose their motivation, as well as interest in things around them, and these effects are reflected in the patients’ speech and facial expressions. But those observations are difficult to quantify.

With recent developments in many technological fields, the collection and analysis of a variety of data sets has become easier and less expensive. For example, a subject’s pause time and speech rate during phone conversations [32], the intensity of a subject’s smile [33], and sensor-detected body motion [34] were used to diagnose and/or assess the severity of subjects’ depression. Observations are not limited only to clinical settings; research using actigraphy to observe subjects’ daily activities has been ongoing for some time, and has been able to find certain differences between depressed patients and healthy volunteers [35].

Methods for automatically detecting and evaluating the severity of neurocognitive disorders include one based on acoustic characteristics [19, 36–40], one using linguistic information [41–43], and some fusing both those approaches [44, 45]. Wearable devices that gather information about activity, sleep, and conversation time were also used to diagnose/assess severity [46].

In many cases, studies that collect large amounts of data from electronic devices also use machine learning to estimate the presence and/or severity of illnesses. When applied to this goal, machine learning approaches are valuable, as data from such applications often contain complex cross-sectional and longitudinal patterns. These complex patterns are exhibited in the joint distribution, and in the linear and nonlinear relationships between all, or subsets of, the aforementioned factors. These relationships are further complicated when utilizing multimodal data. By collecting such data with diagnoses and/or severity information as labels, we can develop novel machine learning techniques to discover these complex patterns, which can in turn provide objective indices and predictive models for diagnosis (categorical classification) and severity assessment (continuous variable prediction), as well as for judging whether there has been an improvement/deterioration in a patient’s condition since their previous visit (categorical classification). Through these machine learning tasks, it is also possible to gain additional insights into which clinical characteristics are helpful in diagnosing and evaluating severity, how to identify characteristics that parallel symptom improvement, and more.

The Project for Objective Measures Using Computational Psychiatry Technology (PROMPT), which is funded by the Japan Agency for Medical Research and Development (AMED), is an industry-academia collaborative research project that aims to develop new techniques for diagnosing and evaluating illness severity utilizing the technology described above; such technology is already readily available or developed by companies in this partnership. In concrete terms, we will record subjects’ facial expressions, speech (acoustic analysis, speech speed analysis, natural language processing), and body motion in an examination room within a simulated clinical visit setting. With subject consent, we will ask them to put on a wearable device to also record daily activity and sleep. The collected data will be combined, machine learning will be applied, and we will develop learning models based on a variety of objectives, with the hope that this research will prove useful in every-day clinical settings and clinical trials. This study was approved by the Institutional Review Board of Keio University School of Medicine and the participating medical facilities. Any adverse events that occur during the study will be reported to and managed by this same review board.

## Methods

### Participants

This study is a multi-site prospective observational study. Participants are recruited at seven hospitals and three outpatient clinics that specialize in treating either mood disorders or dementia, or both, in five different prefectures in Japan. Patient recruitment is conducted in the following locations and hospitals: Tokyo (Keio University Hospital, Tsurugaoka Garden Hospital, Oizumi Hospital, Komagino Hospital); Shiga (Biwako Hospital); Yamagata (Sato Hospital); Fukushima (Asaka Hospital). Outpatient clinics were used for additional patient recruitment in Tokyo (Oizumi Mental Clinic, Asakadai Mental Clinic) and Kanagawa (Nagatsuda Ikoinomori Clinic). Healthy controls were recruited from the same areas as patients. Participants are inpatients or outpatients aged ≥20 years, who met the DSM-5 criteria for major depressive disorder, bipolar disorder, major neurocognitive disorder, and mild neurocognitive disorder. Patients with subjective cognitive impairment (i.e., patients who feel they are cognitively impaired, but when tested, are not shown to have abnormalities) are also included in this study. Exclusion criteria include: (1) paralysis or involuntary movement in the face or body; and (2) inability to speak (e.g., removal of vocal cords). Healthy controls are screened by using the Mini-International Neuropsychiatric Interview (M.I.N.I.) and MMSE, and are excluded if they have a history of psychiatric disorders or show cognitive impairment. Researchers obtain written informed consent from all participants. In cases where patients were judged to be decisionally impaired, the patients’ guardians will give consent. Participants are able to leave the study at any time.

### Assessments

All assessments are undertaken by trained research psychiatrists and/or psychologists. Clinical characteristics (e.g., age, sex, duration of illness), past medical history, and currently prescribed medications are collected using patients’ medical charts. RGB and infrared video recordings [RealSense R200 (Intel Corporation)/ Microsoft Kinect for Windows v2 (Microsoft Corporation)], and voice recordings using an array microphone [Classis RM30W (Beyerdynamic GmbH & Co. KG)/ PRO8HEx Hypercardioid Dynamic Headworn Microphone (Audio-Technica Corporation)], are captured during a 10-minute interview with a psychiatrist and/or psychologist. During the interview, conversations between the interviewer and patient cover topics that arise in normal clinical practice, such as mood, daily living, sleep, events in the past week, concerns, etc. After the 10-minute clinical interview, a semi-structured interview using the clinical assessment tools is conducted (Table 1). In addition to participating in the above-mentioned interview recordings, participants are asked to wear wearable devices [Silmee W20 (TDK Corporation)] until their next assessment. Silmee is a wristband-type wearable monitor equipped with an accelerometer, gyrometer, pulse sensor, thermometer, and UV meter. We make the use of wearable devices optional, as it is possible that some participants will see it as a burden. In order to collect various data from the same patients in different states, assessments are done up to 10 times for each patient during the study period. Visit intervals are not fixed, but we attempt to time them for when patients’ clinical symptoms have changed from the last visit (e.g., if we learn from the treating psychiatrist that a patient has recovered, we attempt to see the patient at that time), so that we can input datasets reflecting various illness severities into the machine learning program. The minimum interval sets are one week for patients with depression and one month for healthy volunteers. The Structural Clinical Interview for DSM-5 (SCID) is performed to the greatest degree feasible to confirm the diagnoses during the follow up period. Normal treatment is continued during the study period. The documents pertaining to this research are only stored in cabinets that lock within a research room of the Keio University School of Medicine’s Department of Neuropsychiatry. We assign research numbers to data that will be used in the study, and from there on, the data are managed using those numbers. Once numbers are assigned, all data are completely separated from any personal identifiers. Additionally, case report forms are managed using electronic data capture.

**Table 1.** A semi-structured interview using clinical assessment tools

| Type of assessment | Time administered | Healthy controls | MDD | BD | Neurocognitive disorder |
| --- | --- | --- | --- | --- | --- |
| HAM-D | Every visit | ✓ | ✓ | ✓ |  |
| MADRS | Every visit | ✓ | ✓ | ✓ |  |
| YMRS | Every visit | ✓ | ✓ | ✓ |  |
| BDI-II | Every visit | ✓ | ✓ | ✓ |  |
| PSQI | Every visit | ✓ | ✓ | ✓ |  |
| MMSE | Screening, every visit | ✓ |  |  | ✓ |
| CDR | Every visit | ✓ |  |  | ✓ |
| LM (Immediate/Delayed) | Every visit | ✓ |  |  | ✓ |
| CDT (copying/free-drawing) | Every visit | ✓ |  |  | ✓ |
| NPI | Every visit | ✓ |  |  | ✓ |
| GDS | Every visit | ✓ |  |  | ✓ |
| M.I.N.I. | Screening | ✓ |  |  | ✓ |
| SCID | Once during the follow up | ✓ | ✓ | ✓ | ✓ |

MDD: Major depressive disorder, BD: Bipolar disorder, HAM-D: Hamilton Depression Rating Scale, MADRS: Montgomery-Asberg Depression Rating Scale, YMRS: Young Mania Rating Scale, BDI-II: Beck Depression Inventory Second Edition, PSQI: Pittsburgh Sleep Quality Index, MMSE: Mini-Mental State Examination, CDR: Clinical Dementia Rating, LM: Wechsler Memory Scale-Revised Logical Memory, CDT: Clock Drawing Test, NPI: Neuropsychiatric Inventory, GDS: Geriatric Depression Scale, M.I.N.I.: Mini-International Neuropsychiatric Interview, SCID: Structural Clinical Interview for DSM-5

All these data are stored securely in Microsoft Azure. Microsoft Azure is a highly reliable cloud-based system, and it has wide compliance with industry-specific and global regulations, such as: adherence to ISO 27001, an international regulation for information security management systems; adherence to FedRAMP, a cloud-computing security standard in the United States; and adherence to ISO/IEC 27018, the international performance standard for regulating how personal information is handled by cloud service providers.

### Analysis

The machine learning models for PROMPT are trained to perform the following tasks: 1) predict whether a subject has or does not have depression/neurocognitive disorders for screening purposes; 2) predict the severity of a subject’s depression/cognitive decline based on results from severity rating scales such as HAM-D (including the 21, 17, and 6 item versions’ scores), MADRS, Beck Depression Inventory, Second Edition (BDI-II), and MMSE with a known margin of error for the predicted rating; 3) predict the improvement or deterioration of a subject’s depressive state/cognitive function with respect to a previously recorded state if the subject has had a prior assessment by the system; and 4) predict the scores of individual items in a depression/cognitive rating scale that are indicators of different aspects of a subject’s depression/cognitive states, such as depressed mood, anhedonia, insomnia, anxiety, and psychomotor retardation/agitation for depression, or orientation to time and place, memory, attention and calculation, language, and visuospatial perception for neurocognitive disorders.

The data used to train these machine learning models are multimodal in nature, including facial expression and eye blinking features extracted from RGB video recordings, body motion features extracted from infrared recordings, and voice features extracted from audio recordings. We first perform data cleaning and feature engineering to construct feature vectors in which the machine learning algorithms can more easily find patterns that can correctly identify healthy and depressed subjects or predict a fine gradient of depression severity from a subject’s physical symptoms.

### Extracted Data

In audio engineering, phonic data are often used to describe the sound generation from the vocal cord and sound modulation from the shape of the mouth and the position of the tongue. To use these physical properties in our machine learning models, we extract phonic data from audio recordings with software such as Praat [47] and openSMILE [48] at 10-ms intervals. These phonic data include: fundamental frequency (F0); first, second, and third formant frequencies (F1, F2, F3); cepstral peak prominence (CPP); and mel-frequency cepstrum coefficients (MFCC).

To discover patterns at a higher level, prosodic speech data are extracted from audio recordings, including: rate of speech, which measures the number of syllables spoken per minute; delay of reply, which measures the length of delay between the end of the physician’s sentence and the beginning of the subject’s subsequent sentence; and pause time, which measures the length of delay between two consecutive sentences spoken by the subject.

Facial features are extracted from video recordings with software such as OKAO Vision and Openface [49, 50]. The data extracted include predicted facial expressions of the subject in each frame of the video recording, and the inverse distance between the upper and lower eye lids.

Regarding body motion, the speed statistics and angles formed by four joints in XYZ dimension, namely Spine Shoulder, Head, Shoulder Right, and Shoulder Left, are utilized as features. These joints are extracted either by Kinect V2 joint map, or from Intel RealSense.

We collect daily activity data for the subjects using wearable devices as described above. Daily activity data targeted for collection include number of steps taken, energy expended, body motion, sleep state, skin temperature, heart rate, and UV exposure index.

### Feature Engineering

For some machine learning models, we need to perform feature engineering to summarize the time-course data extracted from the raw audio and video recordings, and to capture the relationship between pairs of time-course data. The following feature engineering approaches are used to construct features from the multi-modal data as input to the machine learning models for predicting a subject’s depression/cognitive status and/or severity using the following methods: 1) space-delay matrix [51] that computes all pair-wise similarities between the extracted data (space) at each delay from a set of different delay scales (delay); 2) distribution statistics (5-, 25-, 50-, 75-, 95-quantile and mean and standard deviation); 3) Markov transition probabilities for the state change between two adjacent time-series samples; 4) similarity measures between different data; and 5) decision-tree-based quantization of data.

### Machine Learning Architecture

We take two approaches to the machine learning architecture: one based on non-deep-learning machine learning algorithms, utilizing feature selection of the engineered features and meta-models; and one based on deep-learning algorithms.

For the non-deep-learning-based machine learning architecture, we first perform feature selection to choose a subset of the engineered features to build our models. The parameters obtained through feature engineering are passed to an elastic-net model [52] for feature selection. The labels of the dependent variables are regressed on the feature vector and an elastic-net model is fitted. The fitted model has a sparse set of coefficients; i.e., many of the features’ coefficients will be forced to zero during fitting and contribute nothing to the prediction of the labels. The features in the feature vector that have non-zero coefficients are retained as selected features and used to build the next layer of the machine learning algorithm.

Next, the selected features from the elastic-net feature selection layer are used to train the first layer models of the meta-model. Models used in the second layer include algorithms such as Support Vector Regression (SVR) [53], Support Vector Machine (SVM) [54], XGBoost [55], Random Forest (RF) [56], Adaptive Boosting (Adaboost) [57], and Adaptive Bagging (Adabag) [58]. The same selected features (features with non-zero coefficients) are used in each of the machine learning models and the labels predicted by each model are passed as features to the second layer of the meta-model.

For the second layer, we can use an algorithm with logistic regression or SVM for classification, or one with a linear model or SVR for regression. The features for this layer are the predicted labels from the previous machine learning layer, and the true labels are regressed against these predicted labels to train the machine learning model.

For deep-learning-based models, we use deep-learning architectures such as Convolutional Neural Networks (CNN) [59, 60], Gated Convolutional Neural Networks (GCNN) [61], Bayesian Neural Networks (BNN) [62], and Long Short-Term Memory Networks (LSTM) [63]. For these models, the time-course features extracted from the raw video and audio data are used directly as input, instead of the engineered features. It should be noted that for either deep-learning or non-deep-learning-based architectures, the models are not limited to those listed above.

For the improvement/deterioration model we use the non-deep-learning machine learning models, where each input feature vector is constructed from the data of two separate interviews with the same subject. For each of the interviews with the same subject, the feature vector is constructed as described above. To construct the feature vector for the improvement/deterioration model, the feature vector of the prior interview is divided elementwise by the feature vector of the latter interview. This new vector of element-wise ratios of the feature vectors of the two interviews is used as the feature vector for the improvement/deterioration model. The machine learning architecture for the improvement/deterioration model is the same as the model presented above.

### Sample Size

To predict the sample size required for the supervised learning performances, we use learning curves to estimate the number of samples required to reach 90% accuracy for classification tasks. An inverse exponential model is fitted to pairs of sample size and cross-validation accuracy to predict the number of samples necessary. For depression, based on the preliminary data that we collected, we estimated a need for approximately 200 patients and 100 healthy volunteers; for dementia, we estimated a need for 100 patients and 100 healthy volunteers. Assuming an average of three assessments per individual participant, we therefore set a target of 1,500 datasets from 500 participants.

## Discussion

The PROMPT study is unique in its purpose and integrative approach. The main purpose of PROMPT is to develop objective digital biomarkers for the assessment of depression/neurocognitive disorders in the hopes of guiding clinical decision-making in clinical settings. There will be tremendous value in noninvasive and easy-to-use methods that do not put additional burdens on clinical practice, and which can be repeatedly conducted not only in daily clinical settings, but also in clinical trials. In this project, we collect systematically observable features of patients (including facial, speech, and movement expressions) during clinical interviews, as well as daily activity measurements for the time between clinical interviews. We follow and assess each participant up to 10 times longitudinally, so that the machine can learn different severities of the diseases. This approach is also helpful to avoid overfitting in machine learning. Based on the collected datasets, we aim to develop a machine learning model to screen these disorders, assess severity, and reveal whether or not the symptoms improved since the last visit.

Specifically, observable signs of patients, such as facial expression and speech rate, are important characteristics of depressive disorders, but psychomotor disturbances in particular have been considered one of the most fundamental features of depression, especially melancholic depression [30]. They are also one of the diagnostic symptoms of major depressive episodes and manic episodes [64]. Psychomotor disturbances may have predictive value for antidepressant treatments, especially for electroconvulsive therapy [27]. Some rating scales have been developed for psychomotor disturbances, including the CORE measurement [65] and the Motor Agitation and Retardation Scale (MARS) [66]. However, these measurements rely on the subjective judgment of the clinicians, and no reliable and/or validated objective measurement methods for psychomotor disturbances have been developed. Therefore, PROMPT strives to overcome these historical issues. In addition, our model could be used as an assessment tool for psychomotor disturbances, and for distinguishing melancholic depression from heterogeneous DSM-defined major depression. It could also be used for investigating the underlying neurobiology of psychomotor disturbances in collaboration with neuroimaging/neurophysiological measurements in future studies.

For neurocognitive disorders, the importance of early intervention and prevention of disease through the modification of therapy methods is being emphasized more and more. However, examinations such as amyloid PET or cerebrospinal fluid tests are not practical in terms of the invasiveness and cost, as well as the facility equipment requirements. In addition, when performing cognitive assessments at the preclinical stage, it is difficult to distinguish between disease-related changes and changes caused by normal aging, since cognitive impairment is still comparatively minor at that stage. As mentioned previously, learning effects can also be a significant problem when a patient is assessed repeatedly, especially in the early phase of a disorder. It would be highly beneficial if a new approach is developed that can identify high risk patients without these issues.

Challenges of the study are as follows. First, the large variability of the subjects makes it difficult to extract the features that commonly reflect disorder severity. For example, if we learn that one’s conversational response time is slower than a healthy control’s, we still do not know if he/she has psychomotor retardation, because we do not know his/her original speed of speech. But at the same time, psychiatrists can judge if someone has psychomotor retardation even if they do not know what he/she was like before the onset of illness. Psychiatrists most likely gather multimodal information from patients for a comprehensive judgement, and a machine may be able to do the same, as long as it is given the same modalities. Nevertheless, the variability of the samples is the most concerning matter for this study, and though this could be resolved to a certain degree by gathering a larger number of datasets, we may still see the machine learning models’ accuracy hit a ceiling at some point. Second, recruiting severe patients is difficult. As this study does not focus on intervention, recruitment may not be as large a problem in this case, but recruiting severe patients is an inherent difficulty in clinical studies. Imbalanced samples for different severities caused by recruitment difficulties may prohibit the machine learning algorithms from achieving a high prediction accuracy. Third, it is very important to keep inter-rater reliability high when diagnosing and/or assessing patients, as assessment scale scores will be the labels for machine learning. Anticipating this issue, the study team developed educational modules to maintain a high quality of ratings, and the inter-rater reliability will be tested using random sampling during the study period. Finally, since data will be collected from typical clinical settings, the recordings may contain a significant amount of optical and acoustic noise from the background, or due to inconsistent instrument setup. Well-designed preprocessing and data cleaning steps will be important to provide high quality features for the machine learning algorithms.

## Data Availability

Not applicable.

## Abbreviations

UMIN: University Hospital Medical Information Network
UI: uncertainty interval
YLDs: years lived with disability
HAM-D: Hamilton Depression Rating Scale
MADRS: Montgomery-Asberg Depression Rating Scale
PET: positron emission tomography
MRI: magnetic resonance imaging
MCI: mild cognitive impairment
MMSE: Mini-Mental State Examination
MoCA: Montreal Cognitive Assessment
PROMPT: Project for Objective Measures Using Computational Psychiatry Technology
AMED: Japan Agency for Medical Research and Development
DSM-5: Diagnostic and Statistical Manual of Mental Disorders, Fifth Edition
SCID: Structural Clinical Interview for DSM-5
M.I.N.I.: Mini-International Neuropsychiatric Interview
RGB: red, green, blue
UV: ultraviolet
ISO: International Organization for Standardization
FedRAMP: Federal Risk and Authorization Management Program
IEC: International Electrotechnical Commission
BDI-II: Beck Depression Inventory, Second Edition
F0: fundamental frequency
F1, F2, F3: first, second, and third formant frequencies
CPP: cepstral peak prominence
MFCC: mel-frequency cepstrum coefficients
SVR: Support Vector Regression
SVM: Support Vector Machine
RF: Random Forest
Adaboost: Adaptive Boosting
Adabag: Adaptive Bagging
CNN: Convolutional Neural Networks
GCN: Gated Convolutional Neural Networks
BNN: Bayesian Neural Networks
LSTM: Long Short-Term Memory Networks
MARS: Motor Agitation and Retardation Scale
MDD: Major depressive disorder
BD: Bipolar disorder
YMRS: Young Mania Rating Scale
PSQI: Pittsburgh Sleep Quality Index
CDR: Clinical Dementia Rating
LM: Wechsler Memory Scale-Revised Logical Memory
CDT: Clock Drawing Test
NPI: Neuropsychiatric Inventory
GDS: Geriatric Depression Scale

## Declarations

### Ethics approval and consent to participate

This study was approved by the Institutional Review Board of Keio University School of Medicine and the participating medical facilities. Researchers obtain written informed consent from all participants. In cases where patients were judged to be decisionally impaired, the patients’ guardians will give consent. Participants are able to leave the study at any time.

### Consent for publication

Not applicable.

### Availability of data and material

Not applicable.

### Funding

This research is supported by the Japan Agency for Medical Research and Development (AMED) under Grant Number JP18he1102004. The Grant was awarded on Oct. 29, 2015 and ends on Mar. 31, 2019. The funding source did not participate in the design of this study and will not have any hand in the study’s execution, analyses, or submission of results.

Japan Agency for Medical Research and Development (AMED)

20F Yomiuri Shimbun Bldg. 1-7-1 Otemachi, Chiyoda-ku, Tokyo 100-0004 Japan

Tel: +81-3-6870-2200, Fax: +81-3-6870-2241

### Authors’ Contributions

T. Kishimoto, AT, KL, KF, TF, MK, M. Yoshimura, YT, TH, YE, T. Kikuchi, MT, SB, JM, BS, TW, AK, M. Yotsui, HT, YM, KS, YS, MM contributed to the design of the study and writing of the manuscript. All authors have read and approved the manuscript.

## Acknowledgements

We gratefully acknowledge the PROMPT collaborators: Minoru Ko, Hiroaki Miyata, Ruriko Otsuka, Koki Kudo, Kyosuke Sawada, Bun Yamagata, Kanako Ichikura, Yuki Ito, Yuriko Kaise, Satsuki Sakiyama, Ayako Sento, Sayaka Hanashiro, Yuki Momota, Yoshitaka Yamaoka, Fumiya Tsurushima, Mao Yamamoto, Daiki Tsuburai, Kelley Cortright (Keio University), Akiko Goto (Tsurugaoka Garden Hospital), Nobuya Ishida (Biwako Hospital), Yukari Shimanuki, Yuka Oba (Sato Hospital), Inoue Nakamasa (Tokyo Institute of Technology), Kuniko Nishikawa, Akihito Tamiya, Hidefumi Uchiyama, (FRONTEO Healthcare, Inc.), Hiromatsu_Aoki, Haruka Taniguchi (OMRON Corporation), Satoshi Maemoto, Kai Zaremba (SYSTEM FRIEND, INC.), Yasuhiko Fujita, Makoto Hashizume, Koichi Iwase, Kenichiro Shii (Advanced Media, Inc.), Hiroaki Kobayashi (SoftBank Corp.), Nobuki Fujinaka (Microsoft Japan Co., Ltd.), Hideyoshi Murashige (Semco Co.), Fumihiro Kanda (INDUSTRIAL MECHATRONICS CO., LTD).

